# Initial experience with short-course corticosteroids in a small cohort of adults with severe COVID-19 in a tertiary care hospital in India

**DOI:** 10.1101/2020.06.23.20137471

**Authors:** Sanjiv Jha, Kiran Shetty, Sonali Vadi, Sourabh Phadtare, Vatsal Kothari, Abhijit Raut, Sweta Shah, Pallavi Bhargava, Tanu Singhal

## Abstract

Severe COVID 19 disease is associated with high morbidity and mortality with limited therapeutic options. The role of glucocorticoids in treatment of COVID 19 has been riddled with controversy. The study site has been using glucocorticoids in patients with severe COVID 19 since the first few patients of COVID 19 that were admitted. In the initial cohort of 7 patients with severe COVID disease, use of methylprednisolone in a dose of 30 mg twice daily was associated with rapid improvement in oxygenation and decline in CRP levels. While six patients made a complete clinical recovery, one patient died. There were no secondary infections.

---

India is in the midst of the COVID-19 epidemic with steady rise in the numbers of cases and deaths (1). In Mumbai, one of the epicentres of the outbreak several private hospitals have set up COVID treatment units. The study site has set up a negative pressure 17 bed COVID-19 unit since 23rd March 2020. The infection prevention, admission and treatment protocols are in line with recommendations from Government of India (2). Severe COVID 19 is associated with high morbidity and mortality. Treatment options are limited. There has been a controversy about the benefit of steroids in severe COVID 19. We describe here our experience in managing our first few severe cases of COVID-19 with glucocorticoids over a 2 weeks period between 26^th^ March and 8^th^ April 2020. The Institutional Research and Ethics Committee granted a waiver of review for the study.

Eighteen adults with a confirmed diagnosis of COVID-19 by RT-PCR were admitted to the unit in the study period. Of these seven were having severe disease (defined as oxygen saturation ≤93% at any time during the hospital stay), 10 had mild symptoms and one was asymptomatic. Further discussion is confined to the seven patients with severe disease (Table 1). Five were male and the mean age was 58 years (range 44-70 years). Four patients had comorbidities including hypertension (3), diabetes mellitus (2), chronic kidney disease (1) and obstructive airway disease (1). The mean duration of symptoms prior to admission was 7 days (range 4-10 days). Fever, cough and breathing difficulty were present in all patients; one patient had gastrointestinal symptoms of diarrhoea and vomiting. Three were febrile, all had tachycardia but none had hypotension. Five patients were tachypneic and hypoxic at admission while two patients (1 and 6, Table 1) developed hypoxia on day 4 and day 3 of hospitalization. The investigative profile of the study patients at admission is summarized in Table 2. Blood cultures sent at admission were negative in all patients. In the 5 patients who were hypoxic on admission, radiologic investigations were carried out and were abnormal (CXR in 3, CT in 1 and both CXR and CT in 1). The CT scans showed the classic findings of bilateral peripheral consolidations and ground glass opacities. The two patients (1 and 6, Table 1) who became hypoxic later on day 4 and day 3 respectively had abnormal CXR on the day of deterioration.

**Table 1:**
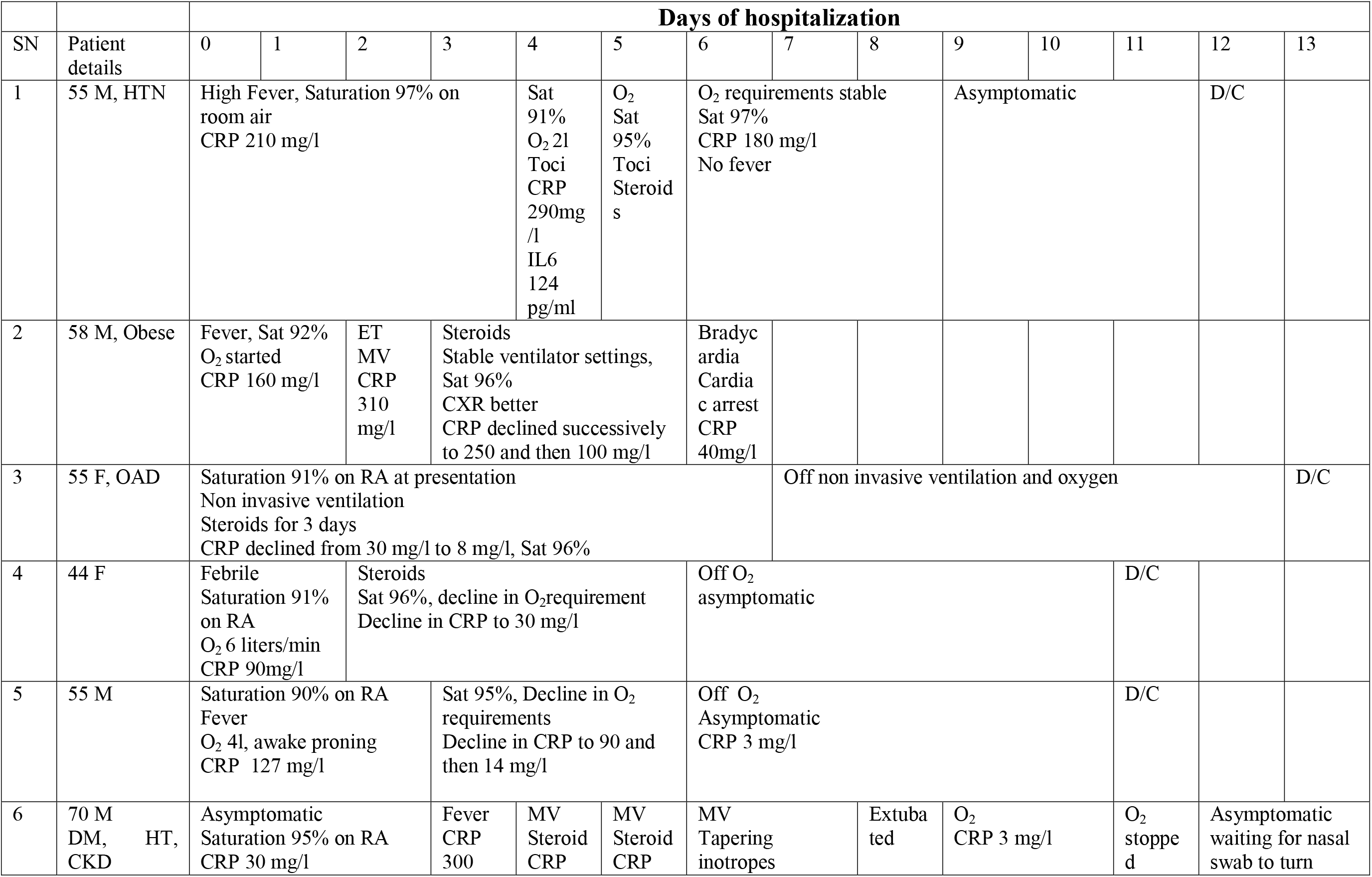
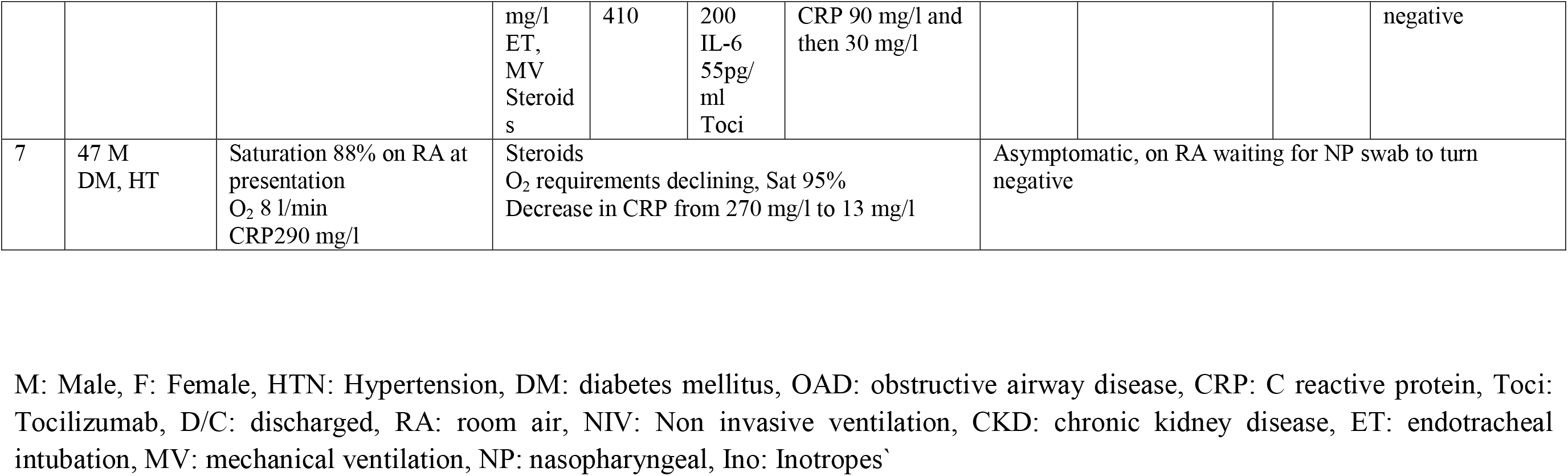
Clinical course of the study patients

**Table 2:** Investigations of study patients at admission

|  | **Absolute lymphocyte count/ mm<sup>3</sup>** | **ANC: ALC** | **Platelet count/mm<sup>3</sup>** | **CRP mg/l** | **PCT ng/ml** | **Creatinine mg/dl** | **AST/ALT IU/l** | **LDH mg/l** | **CPK IU/l** | **D Dimer** | **Ferritin ng/ml** |
| --- | --- | --- | --- | --- | --- | --- | --- | --- | --- | --- | --- |
| Abnormal defined as | <1000 | >3.5 | <150,000 | >6 | >0.5 | >1.4 | >80/100 | >225 | >171 | >500 | >274 |
| Patient 1 | 1100 | 2.45 | 137,000 | 210 | 0.2 | 1.04 | 28/34 | 248 | 110 | 881 | - |
| Patient 2 | 500 | 21 | 255,000 | 160 | 1.91 | 1.08 | 24/21 | 496 | 327 | 643 | - |
| Patient 3 | 1400 | 4 | 331,000 | 30 | 0.3 | 1.38 | 46/16 | 304 | 104 | 1335 | 90 |
| Patient 4 | 1400 | 2.85 | 223,000 | 90 | 0.24 | 0.61 | 184/160 | 715 | 69 | - | - |
| Patient 5 | 1100 | 4 | 259,000 | 127 | 91.6 | 0.86 | 34/32 | 306 | 140 | 804 | 339 |
| Patient 6 | 1800 | 3.3 | 187,000 | 30 | 0.1 | 1.83 | 36/26 | 310 | 185 | 579 | - |
| Patient 7 | 700 | 6.28 | 358,000 | 290 | 0.69 | 1.6 | 71/55 | 427 | 143 | 2006 | 715 |
| **Abnormal** | **3/7** | **4/7** | **1/7** | **7/7** | **3/7** | **2/7** | **1/7** | **7/7** | **3/7** | **6/6** | **2/3** |

As per institutional protocol all these patients were initiated on parenteral antibiotics (ceftriaxone in 6 and piperacillin tazobactam in 1), azithromycin, hydroxychloroquine (400 mg twice daily on day 1 and then 200 mg twice daily), low molecular weight heparin and vitamin C. Intravenous fluids were given as per requirement. Respiratory support during hospitalization included oxygen by nasal prongs/ non rebreathing mask in 4 patients, non invasive ventilation in one and mechanical ventilation (MV) in 2 patients.

The treatment protocol was modified on 30^th^ March 2020 to include corticosteroids after telephonic discussion with a member of the Henry Ford COVID 19 management task force at Detroit, Michigan, USA based on the encouraging results seen by their team with use of intravenous methylprednisolone 30 mg twice daily for 3 days in hospitalized COVID 19 patients. All 7 study patients thus received intravenous methyl prednisolone in a dose of 30 mg twice daily either at admission (patients 3,5) or when they became hypoxic (patients 1 and 6) or when they did not improve with conventional therapy (2,4,7). The response to therapy was monitored by change in clinical parameters including fever, oxygen saturation, requirement of respiratory support and CRP levels. Over the next few days, there was a significant decline in inflammatory markers including CRP, improvement in CXR and reduction of oxygen requirements (Table 1). Steroids were discontinued after 3 days in 6 patients but continued for 7 days in one patient (no 7) who had prolonged hypoxia (Table 1). Tocilizumab was used in 2 patients. In patient no 1, who had persistent fever, rising CRP and who developed hypoxia on day 4, two doses of tocilizumab 400 mg were given first followed by steroids. In patient number 6 a single dose of tocilizumab 400 mg was given 48 hours after initiation of steroids in view of high IL-6 levels and declining but persistently elevated CRP. One patient (No 2) who showed improvement in the ventilation parameters and CXR and decline in CRP after initiation of steroid therapy died suddenly on day 6 following bradycardia and cardiac arrest). Of the other 6 patients, 4 have been discharged home and 2 are in hospital on room air. As per ICMR and state protocol patients can be discharged only when two successive nasopharyngeal swabs turn negative (2). The mean duration of respiratory support required by the study patients was 6 days (range 5-8 days), mean duration to achieve nasopharyngeal swab negativity was 10 days (range 5-12 days) and mean hospital stay in discharged patients 11 days (range 6-13 days). None of the patients developed clinical or culture proven sepsis during the hospital stay.

It is well established that the cytokine storm with excess production of various interleukins (IL1 IL2, IL6, IL7, IL10), GCSF, and TNFα is central to the pathogenesis of COVID-19. Hence it is proposed that use of immunomodulatory therapy including corticosteroids and IL6 inhibitors such as tocilizumab could prove useful (3,4). The literature till some time ago was divided about the benefit of steroids in COVID-19. Most of the international guidelines recommended against use of steroids owing to previous lack of efficacy in severe acute respiratory syndrome (SARS), MERS (Middle East Respiratory syndrome) and pandemic influenza, risk of super infections and increased viral shedding (4,5,6). On the other hand, Chinese experts recommended using low to moderate dose steroids for 5-7 days in patients with moderate to severe ARDS due to COVID 19 (7). A recently published single pre-test, single post-test quasi-experiment by the Henry Ford COVID-19 management task force (on whose advice we started using corticosteroids) have reported significant impact of early short course methyl prednisolone on clinical outcomes, length of stay and mortality in patients with moderate to severe COVID-19 (8). Very recently the recovery trial from the UK reported mortality benefit with use of dexamethasone 6 mg once daily for 10 days (9).

While ours is a small cohort, we have replicated the results from the previous studies in Indian patients. We have also demonstrated good results with methylprednisolone as against dexamethasone used in the Recovery trial. The strength of the study lies in precise documentation of the clinical/ radiologic course and investigative work up of the patients. The limitations of the study include its small sample size, observational nature, single centre experience, absence of a control group and use of other interventions (respiratory support and tocilizumab) which may have contributed to clinical improvement. Also clinicians should remember that patients with COVID 19 can have respiratory deterioration due to a variety of reasons including pulmonary thromboembolism, myocarditis, evolving fibrosis and bacterial infections which will not improve/ may worsen with steroids. Hence careful selection of patients, close monitoring and good supportive care is essential when steroids are administered in COVID 19.

## Data Availability

The raw data is archived with the medical records department of kokilaben hospital

